# Exponential damping: The key to successful containment of COVID-19

**DOI:** 10.1101/2020.03.22.20041111

**Authors:** Feng Zhang, Jinmei Zhang, Menglan Cao, Yong Zhang, Cang Hui

## Abstract

Due to its excessively high capacity for human-to-human transmission, the 2019 novel coronavirus disease (COVID-19), first reported in Wuhan in China, spread rapidly to the entire nation and beyond, and has now been declared a global public health emergency. Understanding the transmission pattern of the virus and the efficacy of transmission control measures is crucial to ensuring regional and global disease control. Here we propose a simple model based on exponential infectious growth, but with a time-varying, largely damping, transmission rate. This model provides an excellent fit to the existing data from the 102 countries and regions which have reported cases for more than 6 days, and, we think, has largely captured the transmission patterns of the COVID-19 outbreak under a variety of intervention and control measures. We found that the damping rate, defined as the rate of the exponential decline in transmission rate, ranged from -0.125 to 0.513 d^-1^ globally (a negative damping rate represents acceleration in spread). The estimated peak time (when the fastest spread occurs) and the final number of infections were found to be greatly affected by the damping rate. Successful control measures, such as those implemented in China and South Korea, have resulted in a clear pattern of exponential damping in the viral spread (also shown during the 2003 outbreak of Severe Acute Respiratory Syndrome, SARS). The damping rate, therefore, could be used as an indicator for the efficacy of implemented control measures. Our model suggests that the COVID-19 outbreak is currently accelerating worldwide, especially rapidly in certain countries (e.g. USA and Australia) where exponential damping is yet to emerge. Consistent with the message from the World Health Organisation (WHO), we thus strongly suggest all countries to take active measures to contain this global pandemic. Slight increments in the damping rate from additional control efforts, especially in countries showing weak or no exponential damping in COVID-19 transmission, could lead to a radically more positive outcome in the fight to contain the pandemic.

## Introduction

The 2019 novel coronavirus (COVID-19), which can cause acute pneumonia, was first reported in Wuhan in December 2019, the capital of Hubei Province in central China (*1, 2*). Due to the excessively high rate of human-to-human transmission, the virus has quickly spread across all provinces of China and all countries of the world (*3*). In order to contain this outbreak, governments and healthcare authorities across the globe have taken a series of strict public health measures. Wuhan and all major cities in Hubei, for instance, were sealed off, human movement and traffic prohibited, quarantine imposed on all potentially exposed people, makeshift hospitals quickly built to receive and cure for infected patients. After implementation for just one month, these control measures effectively contained the spread of this highly infectious novel coronavirus in China (*4*), and were considered therefore highly efficient by the World Health Organization (WHO)(*5*). As the first wave of the pandemic has passed beyond China, COVID-19 now begins to rage worldwide, sweeping across all continents except Antarctica (*6*). For effective monitoring and containment of the pandemic, it is crucial to understand the patterns of its rapidly changing and localised transmission and promptly evaluate whether the currently implemented control measures are adequate to ‘flatten the curve’.

Traditional epidemiological models, such as the SIR and SEIR models, explain the rapid increase in the number of infections by the presence of a large susceptible population exposed to infection, and the decline of infection by the depletion of the susceptible population (*7*). Such a model structure is questionable for capturing the spread of COVID-19 due to the massive size of regional and global susceptible populations (easily running into tens or hundreds of millions of residents in a region). The relatively limited infection, albeit excessively high when focused solely on the sheer number of infections, as well as the resultant mortality, have rather small effects on the demography of regional and global populations, unless a large fraction of the population eventually contracts the virus. In addition, the parameterisation of such models is also unreliable for a novel virus where its pathology and transmission pathways remain unclear with little data support. As such, we here propose a population ecology model with a time-varying infection rate to capture the transmission patterns of COVID-19. The advantage of this phenomenological model is that it does not rely on detailed pathology, yet can still provide an accurate and rapid assessment of COVID-19 transmission patterns under implemented control measures. The rate of exponential damping in transmission rate, as will be shown, provides a real-time evaluation of the efficacy of any implemented control measures.

## Methods and Results

Assuming the population is large yet the outbreak limited, so that its impact on the demographic dynamics of the population itself is negligible, we could capture the number of infected cases *N*(*t*) over time using an ordinary differential equation, *dN*(*t*)/*dt* = *r*(*t*)*N*(*t*)(1− *N*(*t*)/*K*), where *r*(*t*) is the time-dependent transmission rate and *KK* the carrying capacity of the number of infections (set as 70% of the entire population, but please note, in most cases the final number of infections is much lower than *K*, so we have essentially ignored its effect on the outbreak). We estimated the transmission rate as *r*(*t* +1/2) = In*N*(*t* +1) − In(*N*(*t*)), where *tt* is measured in days. Notably, *dr*(*t*)/*dt* > 0 represents the acceleration of the epidemic spread, while *dr*(*t*)/*dt* < 0 the deceleration and damping dynamics. We define the damping rate (*a*) as the rate of the exponential decline in the transmission rate *r*(*t*); that is, *r*(*t*) = *e*^−*at*+*b*^. An effective control measure should, arguably, result in the deceleration of the spread at a high damping rate (large positive *a*), while inadequate control measures could lead to a low damping rate (small positive *a* close to zero) and even the acceleration of the spread (*a* < 0). The solution to the above differential equation is 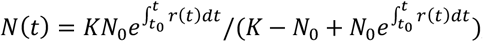, where *N*_0_ is the number of infected cases at the initial time (*t*_0_), in particular 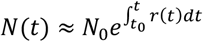 when ignoring *K*’s effect. Thus, to contain the virus outbreak with any measures, it is necessary to ensure the convergence of 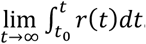. The peak of the outbreak happens when the infection increases at the fastest pace (*d*^2^*N* (*t*)/*dt*^2^ = 0) and can be calculated by solving the equation *dr*(*t*)/*dt* +*r*(*t*)^2^(1− 2*N*(*t*)/*K*) = 0 (appr. *dr*(*t*)/*dt* +*r*(*t*)^2^ = 0). In this simple model, the transmission pattern of an outbreak can be captured solely by the transmission rate *r*(*t*) itself, which reflects the compound effects of the natural transmission rate under implemented control measures.

To illustrate our model, we first compiled the daily numbers of COVID-19 infections from the website of the NHCC (www.nhc.gov.cn) for the period of 10 January to 3 March 2020 in Wuhan city, Hubei Province, and the whole of China. Evidently, our model provided an excellent fit to the data, unveiling a clear pattern of COVID-19 transmission (Fig.1). The transmission rates in Wuhan, Hubei (but excluding Wuhan), and the rest of China outside Hubei, all began to decline exponentially at around the same damping rate (about 0.16 d^-1^) after the large-scale control measures implemented by the Chinese authorities from 23 January (red lines in the left panels of Fig.1). Exponential damping was more obvious outside Wuhan after 12 February (at a rate of 0.32 d^-1^; see the blue lines in the left panels of Fig.1). Such exponentially damping patterns have accurately captured the spreading dynamics of COVID-19 in China (see right panels of Fig.1), and thus could be considered a reliable monitoring indicator of the effectiveness of those control measures implemented in other global regions for controlling the COVID-19 outbreak.

**Fig.1:**
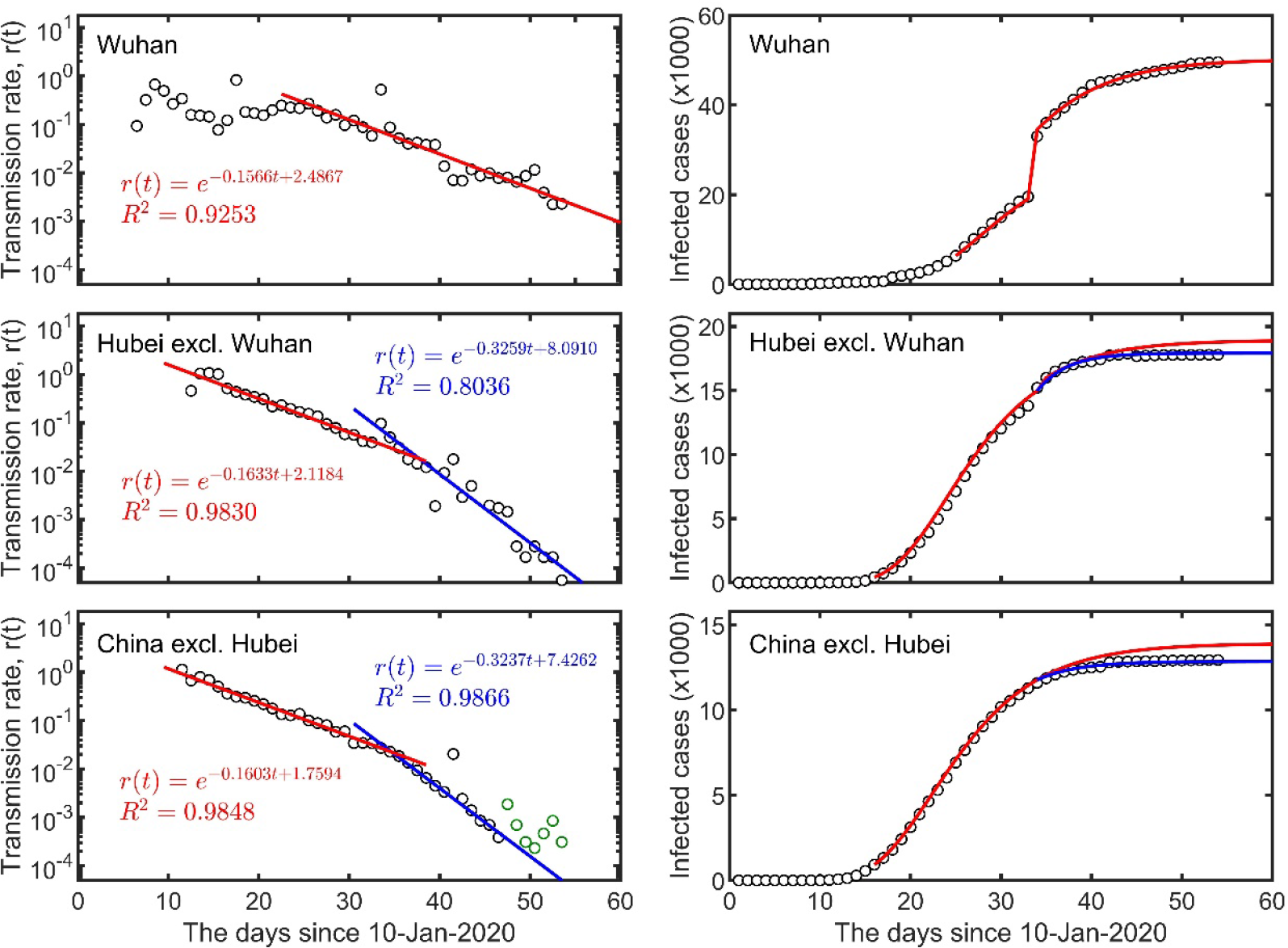
The exponential damping of COVID-19 transmission rates in Wuhan city, Hubei Province (excluding Wuhan) and the rest of China (excluding Hubei) (left column), and the total number of infection cases (right column). The red lines on the left panel represent regressions of the data after 3-Feb-2020 for Wuhan city, from 25-Jan-2020 to 11-Feb-2020 for Hubei province (excluding Wuhan) and the rest of China (excluding Hubei), while blue lines are from regressions based on the data after 12-Feb-2020. The lines on the right panels are the corresponding predictions using the fitted time-dependent transmission rate (*r*(*t*)). Circles indicate real data, and green circles indicate cases imported from other countries but were not considered in the regression.

Using the daily infection numbers from 20 January to 16 March 2020, from the WHO website (www.who.int), we analysed the dynamics of the COVID-19 outbreak in the three worst affected countries (South Korea, Iran and Italy). We found that the transmission rates of COVID-19 in these three countries were all declining exponentially over time, with South Korea experiencing the highest damping rate, similar to that of China (around 0.16 d^-1^), while the other two countries lagged behind at a much lower damping rate, especially Italy (Fig.2). Using the current exponential function of the transmission rate, we estimated the peak of COVID-19 spread at 18 March 2020 (95% CI: 17-26 March) in Iran, and 6 April 2020 (95% CI: 24 March-10 May) in Italy, while the peak in South Korea has passed at 1 March 2020. According to our model, the final number of infected cases in South Korea could reach up to 8,895 (95% CI: 8,593-9,377), in Iran 42,781 (95% CI: 28844-87335), and in Italy 370,841 (95% CI: 116,538-617,355).

**Fig.2:**
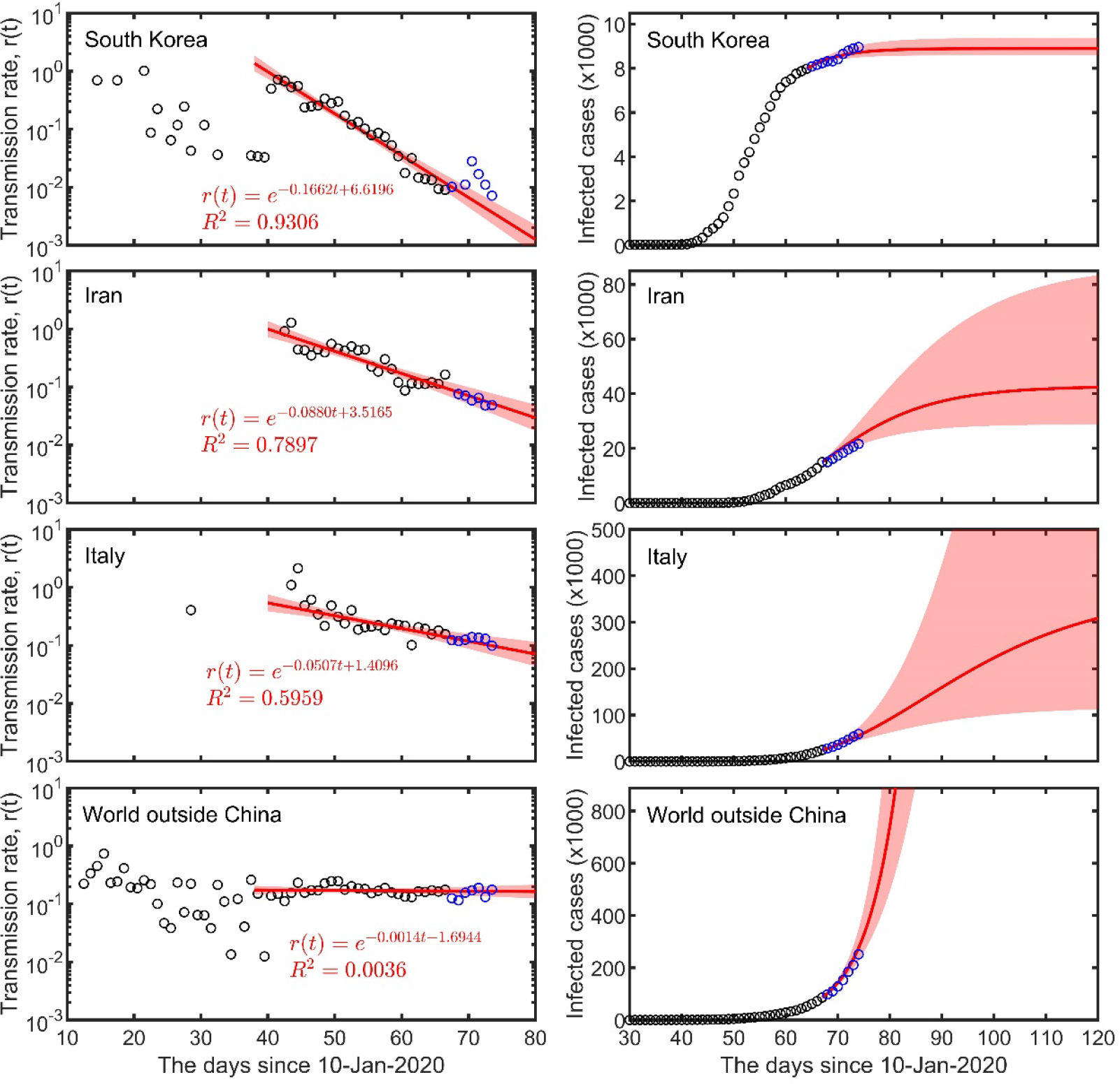
The transmission and spread of COVID-19 outside China based on reported cases. The red lines on the left panel represent regressions of the data from 18-Feb-2020 for South Korea, from 20-Feb-2020 for Iran, from 2-Mar-2020 for Italy, and from 18-Feb-2020 for the entire world except China, to 16-Mar-2020. The red lines on the right panel are predictions based on the corresponding regressions on the left panel. Shadows indicate 95% confidence intervals. The data before 16 March 2020 (used for regression), are shown as black open circles, and after this date as blue open circles.

We further calculated the damping rate for all 102 countries and regions that have reported 7 or more days of infection, from which we also estimated the peak time since first local infection, and the final number of infections (Table S1). Results suggest a large variation in the damping rate of COVID-19 transmission across the world (Fig.3a), from effective control (*a*>0.14 d^-1^) in 14 countries, to outbreak acceleration (*a*<-0.01 d^-1^) in India, USA, Canada, Australia, Singapore and Thailand (Table S1).

**Fig.3:**
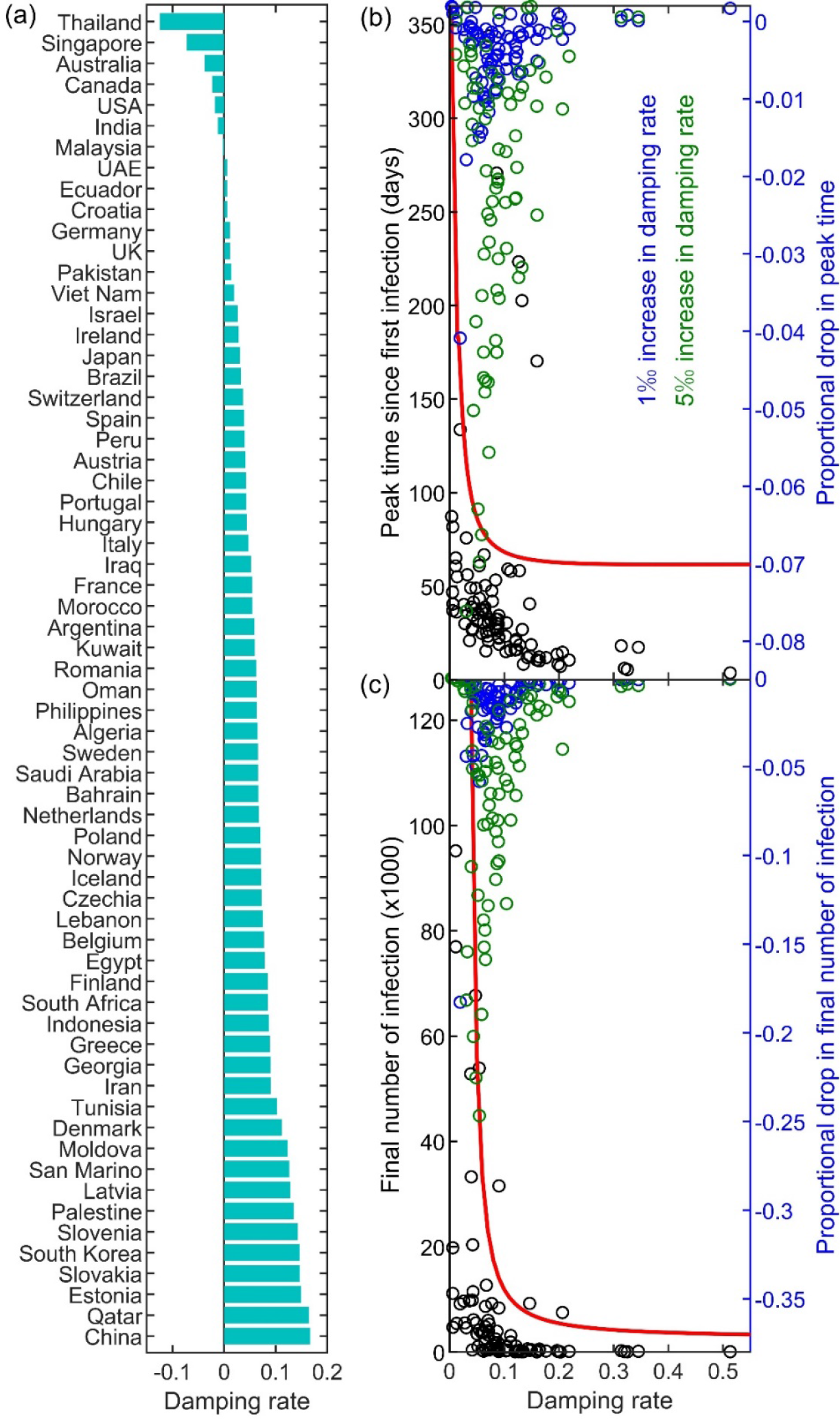
The damping rate and the spread of COVID-19 by country (or region). Estimated damping rate (a) ranked from low to high for 64 countries with more than 12 calculated transmission rates available for regression until 24 March 2020. (b): The relationship between estimated damping rate and the peak time since first infection (reported confirmed case) and (c): between estimated damping rate and the final number of infections for 100 countries (or regions) with more than 6 transmission until 24 March 2020. Estimated values are presented in black circles. The red curves are estimated peak time in (b) and final number of infection (left vertical axis × 100) in (c) for the entire world excluding China if the specific damping rate (as given along the horizontal axis) is implemented from 24-March-2020. Blue and green circles indicate the proportional drop in the peak time since first infection (b) and the final number of infections (c) respectively from a 1‰ and 5‰ increase in the observed damping rate (Table S1).

For 100 countries with more than 6 days of infection, we assessed the effect of 1‰ and 5‰ increments in their observed damping rates on reducing the peak time and the final number of infections. There were strong negative relationships between the damping rate and the peak time since first infection (black open circles in Fig.3b), and between the damping rate and the final number of infections (black open circles in Fig.3c). In countries with low damping rates, the slight increase in the damping rate by 1‰ or 5‰ could drastically reduce the peak time (blue and green open circles, respectively, in Fig.3b) and the final number of infections (blue and green open circles, respectively, in Fig.3c). In countries with high damping rates, such minor increments in the damping rate have trivial effects (Fig.3b-c).

For the entire world outside China, the spread of COVID-19 has been increasing exponentially at a constant transmission rate (0.17 d^-1^) with its damping pattern yet to emerge (the damping rate is close to zero; see the two bottom panels in Fig.2). The pandemic (excluding China) can only be contained if global control efforts, especially with extra efforts in countries showing no clear signs of exponential damping, can suppress the transmission rate *r*(*t*) to decline by a sufficient damping rate (see red lines in Fig.3b-c). To contain the disease by the end of 2020 (one new case every 10 days) in the world (excluding China), we need to reduce the current transmission rate with a clear exponential damping rate of *a*=0.057 d^-1^, with the estimated peak time in 11 April 2020 and the final number of infections of 3,829,338 people.

## Discussion

The data from China and South Korea show that it is possible to contain the spread of COVID-19; this is signalled by the exponential damping of the transmission rate (Fig.1; the two top panels of Fig.2). Such exponential damping is also evident from the data of the 2003 SARS outbreak in mainland China, Hong Kong and the entire world (data from the website of WHO, Fig.S1). This implies that exponential damping in disease transmission could be a universal pattern of successful infectious disease containment. The damping rate of virus transmission reflects the effectiveness of implemented control measures over the natural infection rate of the disease, and its variation across countries therefore reveals whether the current implemented local/regional measures are adequate (see Fig.1, Fig.2 & Fig.3). By estimating the time-varying transmission rate and its damping rate, our model provides a simple theoretical framework for monitoring the spread of an outbreak and assessing the efficacy of implemented control measures in real time. This is important for regional decision-makers and global governance to reflect upon, in order to modify any implemented control measures and practices in time.

Our analysis shows that, at the moment, the pandemic is accelerating exponentially around the world (Fig.2) and an overall damping pattern is yet to emerge. Theoretically, our model can be used for rapid evaluation of the pandemic outbreak in real time and assessment of any intervention measures. To this end, control measures from countries and regions that have already shown exponential damping in their transmission rates could be communicated and compared by the WHO for better local disease control worldwide. Additional control measures should be implemented in countries showing no signs of exponential damping; however, slight improvement of the current control measures can bring about drastic improvement on outbreak control in countries already showing weak exponential damping in transmission. Globally, our analysis suggests that an additional control effort to bring a minimum damping rate of 0.057 d^-1^ is needed to effectively control this COVID-19 pandemic below 4 million infections within 2020.

## Data Availability

All data are accessible to the public through the websites of NHCC (www.nhc.gov.cn) and WHO (www.who.int).

https://www.who.int

## Acknowledgement

FZ is supported by the National Natural Science Foundation of China (No. 31360104) and AHU (No. S020118002/101); CH is supported by the National Research Foundation of South Africa (grant 89967).

## Supplementary File

**Table S1:** Global rankings of the damping rate and its lower and upper bounds of 95% confidence interval (a, LB and UB, respectively), peak time, the estimated final number of infections (FNI), and the number of daily transmission rates (n) used in the model fitting. Estimated for 102 countries and regions with at least 6 days of transmission until 24 March 2020, with K representing 70% of its total population. With 1‰ and 5‰ increase in the observed damping rate, the peak time is expected to shift by PTS days (+1 indicates a delay of 1 day), while the final number of infections is expected to decline by a number of FNID people.

| Country | $a$ | LB | UP | Peak time | FNI | $n$ | PTS 1‰ | FNID 1‰ | PTS 5‰ | FNID 5‰ |
| --- | --- | --- | --- | --- | --- | --- | --- | --- | --- | --- |
| Kazakhstan | 0.5131 | -0.0989 | 1.125 | 19-Mar | 64 | 6 | 0 | 0 | 0 | 0 |
| Jordan | 0.3452 | -0.0024 | 0.6928 | 20-Mar | 151 | 7 | 0 | 0 | 0 | 1 |
| Guadeloupe | 0.3251 | 0.1773 | 0.4729 | 20-Mar | 73 | 7 | 0 | 0 | 0 | 0 |
| Brunei Darussalam | 0.3198 | 0.164 | 0.4756 | 17-Mar | 97 | 10 | 0 | 0 | 0 | 0 |
| Armenia | 0.3138 | 0.2099 | 0.4178 | 20-Mar | 263 | 8 | 0 | 0 | 0 | 1 |
| Burkina Faso | 0.2186 | -0.1627 | 0.5998 | 22-Mar | 136 | 7 | 0 | 0 | 0 | 2 |
| Turkey | 0.2064 | -0.0127 | 0.4256 | 27-Mar | 8,053 | 6 | 0 | 72 | 0 | 349 |
| Jamaica | 0.2027 | -0.0633 | 0.4687 | 18-Mar | 26 | 8 | 0 | 0 | 0 | 0 |
| Albania | 0.1985 | 0.061 | 0.3361 | 18-Mar | 117 | 11 | 0 | 0 | 0 | 1 |
| Serbia | 0.1967 | 0.0944 | 0.2989 | 20-Mar | 291 | 11 | 0 | 1 | 0 | 3 |
| Cyprus | 0.1764 | 0.091 | 0.2618 | 23-Mar | 205 | 8 | -1 | 1 | -1 | 4 |
| China | 0.167 | 0.1443 | 0.1896 | 5-Feb | 81,062 | 29 | 0 | 5 | 1 | 24 |
| Qatar | 0.1642 | 0.0756 | 0.2528 | 12-Mar | 568 | 17 | 0 | 0 | 0 | 2 |
| Bulgaria | 0.164 | 0.0053 | 0.3228 | 20-Mar | 325 | 11 | 0 | 1 | 0 | 5 |
| Andorra | 0.1599 | -0.1622 | 0.482 | 26-Mar | 452 | 7 | 0 | 4 | 0 | 19 |
| Bolivia | 0.1596 | -0.1079 | 0.427 | 22-Mar | 48 | 7 | 0 | 0 | 0 | 1 |
| Estonia | 0.1492 | 0.0518 | 0.2467 | 18-Mar | 503 | 12 | 0 | 1 | 0 | 7 |
| Slovakia | 0.1467 | 0.0834 | 0.2099 | 20-Mar | 327 | 15 | 0 | 1 | 0 | 6 |
| South Korea | 0.1464 | 0.1302 | 0.1627 | 1-Mar | 9,298 | 33 | 1 | 2 | 1 | 11 |
| Slovenia | 0.1424 | 0.0759 | 0.2089 | 18-Mar | 647 | 16 | 0 | 2 | 0 | 10 |
| Palestine | 0.135 | 0.0545 | 0.2154 | 13-Mar | 75 | 15 | 1 | 0 | 1 | 1 |
| Afghanistan | 0.1331 | -0.0513 | 0.3175 | 23-Mar | 96 | 8 | 0 | 1 | 0 | 3 |
| Faroe Islands | 0.1324 | 0.0024 | 0.2623 | 25-Mar | 352 | 10 | 0 | 3 | 0 | 14 |
| Latvia | 0.1282 | 0.0191 | 0.2373 | 22-Mar | 314 | 13 | 0 | 2 | 0 | 9 |
| Sri Lanka | 0.1276 | 0.074 | 0.1812 | 26-Mar | 320 | 10 | 0 | 3 | 0 | 16 |
| San Marino | 0.1263 | 0.0389 | 0.2137 | 15-Mar | 211 | 15 | 0 | 1 | 0 | 3 |
| Moldova | 0.1231 | -0.0134 | 0.2596 | 25-Mar | 273 | 12 | -1 | 2 | -1 | 11 |
| Colombia | 0.1215 | 0.0316 | 0.2115 | 29-Mar | 1,146 | 9 | 0 | 16 | -1 | 77 |
| Senegal | 0.1209 | -0.1816 | 0.4234 | 24-Mar | 175 | 8 | 0 | 1 | -1 | 7 |
| Paraguay | 0.1199 | -0.0553 | 0.2951 | 25-Mar | 69 | 8 | 0 | 1 | 0 | 3 |
| Russian | 0.1119 | 0.0488 | 0.1751 | 30-Mar | 3,196 | 9 | 0 | 56 | 0 | 260 |
| Denmark | 0.1118 | 0.0616 | 0.1619 | 16-Mar | 2,105 | 24 | 0 | 8 | 0 | 37 |
| Cambodia | 0.1067 | -0.0106 | 0.224 | 27-Mar | 350 | 11 | 0 | 5 | 0 | 22 |
| Ukraine | 0.1042 | -0.1821 | 0.3906 | 4-Apr | 967 | 6 | -1 | 27 | -1 | 125 |
| Malta | 0.1031 | 0.0322 | 0.1739 | 27-Mar | 348 | 11 | 0 | 4 | -1 | 21 |
| Tunisia | 0.1024 | -0.004 | 0.2089 | 27-Mar | 273 | 12 | -1 | 3 | -1 | 16 |
| Costa Rica | 0.0906 | -0.0966 | 0.2778 | 28-Mar | 474 | 9 | 0 | 7 | -1 | 33 |
| Iran | 0.0903 | 0.0781 | 0.1026 | 17-Mar | 36,527 | 31 | 0 | 209 | 0 | 989 |
| Dominican | 0.0901 | -0.123 | 0.3033 | 13-Apr | 26,247 | 4 | -1 | 1,640 | -2 | 6,981 |
| Georgia | 0.0899 | 0.0334 | 0.1464 | 20-Mar | 112 | 15 | 0 | 1 | 0 | 4 |
| Azerbaijan | 0.0893 | -0.0203 | 0.1989 | 31-Mar | 430 | 8 | 0 | 9 | -1 | 41 |
| Greece | 0.0886 | 0.0275 | 0.1497 | 25-Mar | 1,949 | 15 | 0 | 25 | 0 | 115 |
| North Macedonia | 0.0884 | 0.0308 | 0.146 | 2-Apr | 1,136 | 11 | 0 | 29 | -1 | 132 |
| Indonesia | 0.0867 | 0.0337 | 0.1396 | 1-Apr | 3,993 | 13 | 0 | 92 | -1 | 422 |
| Luxembourg | 0.0863 | 0.0386 | 0.134 | 7-Apr | 23,191 | 11 | 0 | 846 | -2 | 3,787 |
| Lithuania | 0.085 | -0.0907 | 0.2606 | 4-Apr | 1,908 | 11 | 0 | 57 | -1 | 256 |
| South Africa | 0.0845 | 0.0247 | 0.1443 | 3-Apr | 2,563 | 13 | -1 | 66 | -2 | 301 |
| Finland | 0.0843 | 0.0176 | 0.151 | 26-Mar | 2,011 | 17 | 0 | 27 | -1 | 127 |
| Egypt | 0.0784 | -0.0016 | 0.1584 | 28-Mar | 1,310 | 14 | 0 | 23 | -1 | 105 |
| Belgium | 0.0772 | 0.0186 | 0.1358 | 1-Apr | 22,792 | 17 | 0 | 547 | -1 | 2,487 |
| Lebanon | 0.075 | 0.0357 | 0.1143 | 26-Mar | 812 | 19 | 0 | 13 | -1 | 58 |
| Czechia | 0.0725 | 0.032 | 0.113 | 2-Apr | 8,429 | 18 | 0 | 224 | -1 | 1,009 |
| Iceland | 0.072 | 0.0161 | 0.1279 | 3-Apr | 4,174 | 14 | -1 | 111 | -2 | 503 |
| Norway | 0.0711 | 0.041 | 0.1013 | 27-Mar | 7,679 | 23 | 0 | 135 | -1 | 619 |
| Poland | 0.0705 | 0.003 | 0.1381 | 5-Apr | 6,879 | 16 | 0 | 225 | -1 | 1,004 |
| Panama | 0.0699 | -0.0582 | 0.1981 | 12-Apr | 9,633 | 10 | -1 | 485 | -3 | 2,090 |
| Netherlands | 0.0673 | 0.0475 | 0.0871 | 3-Apr | 28,572 | 24 | -1 | 790 | -2 | 3,543 |
| Bahrain | 0.0665 | 0.0012 | 0.1318 | 15-Mar | 579 | 22 | 0 | 5 | -1 | 22 |
| Saudi Arabia | 0.0657 | -0.0235 | 0.1549 | 10-Apr | 11,013 | 13 | 0 | 495 | -2 | 2,148 |
| Sweden | 0.0654 | 0.0256 | 0.1053 | 28-Mar | 6,909 | 26 | 0 | 133 | -1 | 604 |
| Algeria | 0.0648 | -0.0256 | 0.1552 | 5-Apr | 1,838 | 13 | 0 | 61 | -2 | 269 |
| Philippines | 0.0632 | -0.0229 | 0.1494 | 6-Apr | 3,827 | 14 | 0 | 135 | -2 | 596 |
| Oman | 0.0624 | 0.0111 | 0.1136 | 27-Mar | 183 | 15 | 0 | 3 | -1 | 16 |
| Romania | 0.0622 | 0.0126 | 0.1119 | 7-Apr | 4,539 | 17 | -1 | 166 | -2 | 728 |
| Kuwait | 0.0592 | 0.0011 | 0.1173 | 21-Mar | 436 | 19 | 0 | 6 | -1 | 27 |
| Argentina | 0.0587 | -0.0147 | 0.1322 | 11-Apr | 3,957 | 15 | -1 | 185 | -3 | 797 |
| Morocco | 0.0546 | -0.0327 | 0.1419 | 17-Apr | 4,666 | 12 | -1 | 301 | -4 | 1,246 |
| France | 0.0544 | 0.0336 | 0.0752 | 8-Apr | 154,480 | 25 | 0 | 6,218 | -3 | 26,937 |
| Iraq | 0.052 | 0.0103 | 0.0938 | 1-Apr | 1,069 | 22 | 0 | 30 | -2 | 134 |
| Bosnia-Herzegovina | 0.0505 | -0.1738 | 0.2747 | 27-Apr | 38,813 | 8 | -1 | 4,045 | -4 | 15,554 |
| Belarus | 0.0484 | -0.0352 | 0.132 | 26-Apr | 11,239 | 7 | -1 | 1,080 | -5 | 4,196 |
| Italy | 0.0473 | 0.0361 | 0.0586 | 9-Apr | 513,246 | 29 | -1 | 22,282 | -3 | 95,195 |
| Hungary | 0.0438 | -0.0104 | 0.098 | 27-Apr | 13,572 | 14 | -1 | 1,268 | -6 | 4,921 |
| Portugal | 0.0429 | -0.0041 | 0.0899 | 3-May | 557,445 | 15 | -1 | 65,651 | -6 | 246,241 |
| Chile | 0.0423 | -0.0245 | 0.1091 | 6-May | 340,121 | 14 | -1 | 45,115 | -7 | 163,404 |
| Martinique | 0.0415 | -0.1703 | 0.2533 | 3-May | 6,653 | 8 | -1 | 770 | -7 | 2,872 |
| Austria | 0.0414 | 0.0112 | 0.0716 | 25-Apr | 176,508 | 24 | -1 | 15,151 | -5 | 59,565 |
| Peru | 0.0398 | -0.0556 | 0.1351 | 11-May | 318,960 | 12 | -1 | 49,216 | -8 | 170,614 |
| Spain | 0.0387 | 0.0151 | 0.0623 | 1-May | 2,697,074 | 24 | -1 | 274,860 | -7 | 1,053,324 |
| Switzerland | 0.0366 | 0.0115 | 0.0617 | 4-May | 880,936 | 22 | -2 | 95,284 | -7 | 363,469 |
| Brazil | 0.0326 | -0.0074 | 0.0725 | 26-May | 2,110,518 | 16 | -3 | 431,132 | -12 | 1,353,153 |
| Japan | 0.0307 | 0.0182 | 0.0432 | 6-Apr | 4,746 | 38 | -2 | 216 | -6 | 885 |
| Ireland | 0.0281 | -0.0447 | 0.101 | 26-May | 888,122 | 18 | -2 | 152,343 | -11 | 525,495 |
| Israel | 0.0262 | 0.0028 | 0.0495 | 31-May | 2,384,550 | 21 | -2 | 417,057 | -10 | 1,482,268 |
| Viet Nam | 0.0195 | -0.0605 | 0.0995 | 9-Jun | 9,980 | 15 | -6 | 1,957 | -25 | 5,978 |
| Pakistan | 0.014 | -0.0771 | 0.105 | 20-Jun | K | 15 | 4 | 3,358,141 | 20 | 70,368,628 |
| New Zealand | 0.013 | -0.0458 | 0.0717 | 28-Apr | K | 10 | 0 | 0 | 3 | 129 |
| UK | 0.0114 | -0.012 | 0.0348 | 26-May | K | 26 | 2 | 45,129 | 12 | 2,512,047 |
| Germany | 0.0112 | -0.0264 | 0.0487 | 14-May | K | 26 | 1 | 12,415 | 7 | 790,499 |
| Croatia | 0.0062 | -0.0356 | 0.048 | 12-May | K | 17 | 1 | 0 | 7 | 105 |
| Ecuador | 0.0059 | -0.0894 | 0.1012 | 6-May | K | 14 | 1 | 0 | 5 | 12 |
| UAE | 0.0058 | -0.0114 | 0.0231 | 10-Jun | K | 17 | 4 | 20 | 22 | 59,424 |
| Mexico | 0.0055 | -0.1005 | 0.1115 | 13-May | K | 11 | 1 | 0 | 7 | 31 |
| Malaysia | -0.0022 | -0.0553 | 0.0509 | 8-May | K | 16 | 1 | 0 | 6 | 0 |
| India | -0.0122 | -0.1112 | 0.0868 | 24-May | K | 19 | 2 | 0 | 9 | 0 |
| USA | -0.018 | -0.084 | 0.0481 | 15-Apr | K | 20 | 1 | 0 | 2 | 0 |
| Reunion | -0.0211 | -0.1488 | 0.1066 | 12-Apr | K | 8 | 0 | 0 | 1 | 0 |
| Canada | -0.0229 | -0.047 | 0.0011 | 19-Apr | K | 23 | 1 | 0 | 2 | 0 |
| Australia | -0.0376 | -0.0654 | -0.0098 | 17-Apr | K | 27 | 0 | 0 | 1 | 0 |
| Singapore | -0.0725 | -0.0947 | -0.0502 | 17-Apr | K | 26 | 1 | 0 | 1 | 0 |
| Thailand | -0.1247 | -0.1694 | -0.0799 | 4-Apr | K | 16 | 0 | 0 | 1 | 0 |
| World excl. China | 0.0039 | -0.003 | 0.0107 | 5-Jun | K | 34 | 3 | 92 | 18 | 4,278,152 |

**Fig.S1:**
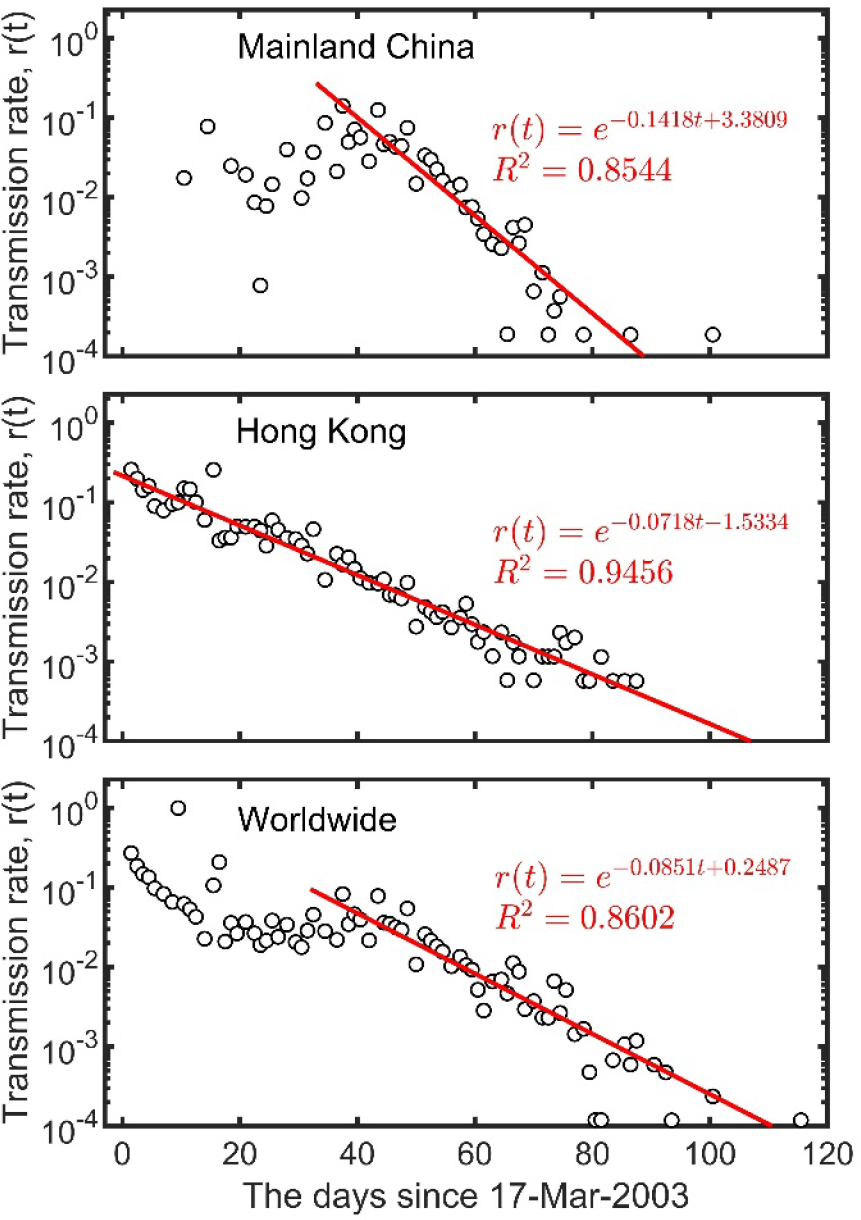
The exponential damping of the 2003 SARS transmission rate in China. Data from the website of WHO (www.who.int).

